# Pneumonia in patients with rheumatoid arthritis: effect of microbial airway colonization

**DOI:** 10.1101/2021.03.15.21253562

**Authors:** Shuhei Ideguchi, Kazuko Yamamoto, Masahiro Tahara, Tomohiro Koga, Shotaro Ide, Tatsuro Hirayama, Takahiro Takazono, Yoshifumi Imamura, Taiga Miyazaki, Noriho Sakamoto, Shimpei Morimoto, Koichi Izumikawa, Katsunori Yanagihara, Kazuto Ashizawa, Takatoshi Aoki, Atsushi Kawakami, Kazuhiro Yatera, Hiroshi Mukae

## Abstract

Rheumatoid arthritis frequently complicates airway diseases and is associated with high rates of pneumonia and mortality. The relationship between microorganisms colonizing the lower respiratory tract and the subsequent incidence of pneumonia in patients with rheumatoid arthritis is unclear. In this study, we aimed to identify microorganisms colonizing the lower respiratory tract in patients with rheumatoid arthritis related to the risk of developing subsequent pneumonia in these patients.

A retrospective cohort study was designed to include a total of 121 patients with rheumatoid arthritis (median age, 67 years; women, 78.5%) who underwent bronchoscopy at three hospitals from January 2008 to December 2017. The following information was extracted from their electronic medical records: patient characteristics, microorganisms detected by bronchoscopy, and subsequent incidences of pneumonia. The patients were divided into groups based on the microorganisms isolated from the lower respiratory tract and compared with control subjects. The cumulative incidence of pneumonia was assessed using the Kaplan–Meier method and log-rank test. Risk factors for pneumonia were analyzed using the Cox proportional-hazards analysis.

The most frequently isolated microbes from the lower respiratory tract in descending order were *Pseudomonas aeruginosa, Staphylococcus aureus*, and *Haemophilus influenzae*. The annual incidence rates of pneumonia per 1000 patients were 100, 62, 132, and 38, in the *P. aeruginosa, S. aureus, H. influenzae*, and control group, respectively. Patients colonized with *P. aeruginosa* had a higher frequency of macrolide use and a higher degree of bronchiectasis than patients in the control group. The rate of the subsequent incidence of pneumonia was higher in the *P. aeruginosa* group (*P* = 0.038), and *P. aeruginosa* was an independent risk factor for pneumonia (hazard ratio, 3.504; 95% confidence interval, 1.153–10.330).

The colonization of the lower respiratory tract by *P. aeruginosa* in patients with rheumatoid arthritis was associated with the subsequent incidence of pneumonia.

## Introduction

The high incidence of pneumonia and associated mortality in patients with rheumatoid arthritis (RA) is a critical concern [1]. Airway diseases (ADs) such as bronchiectasis are common complications of RA, with a rate of 10–30% [2, 3]; patients with RA and ADs have a significantly poor prognosis [4]. *Pseudomonas aeruginosa* increases the risk of mortality and exacerbation in patients with bronchiectasis [4, 5]; however, the relationship between airway colonization and the incidence of pneumonia in patients with RA has not yet been clarified. Thus, we aimed to identify whether colonization of the lower respiratory tract (LRT) by microorganisms, particularly *P. aeruginosa*, correlated with the occurrence of pneumonia in patients with RA.

## Materials and methods

### Patients and study design

This retrospective cohort study was conducted at the Nagasaki University Hospital, University of Occupational and Environmental Health, Japan, and Isahaya General Hospital, from January 2008 to December 2017. This multicenter study was conducted in compliance with the Declaration of Helsinki and was approved by the Ethics Committees of the participating institutions (Nagasaki University Hospital, Approval Number: 19061714; University of Occupational and Environmental Health, Japan, Approval Number: 19-044; Isahaya General Hospital, Approval Number: 2020-9). The requirement for patient consent was waived due to the retrospective nature of the study, which ensured anonymity.

The inclusion criteria were as follows: age ≥20 years, an RA diagnosis by rheumatologists certified by the Japan College of Rheumatology [6], and bronchoscopy performed by a pulmonary physician. We excluded patients if they had active infections requiring antimicrobial agents at the time of bronchoscopy; malignancy diagnosed within five years of the study; tracheotomy; cystic fibrosis (CF); or at least one of the following conditions: human immunodeficiency virus infection, a CD4 cell count <350/μL, solid organ transplantation, or neutropenia <500 cells/μL. Patient characteristics and RA status were extracted from the hospital’s electronic medical records.

### Evaluation of bronchiectasis

Chest computed tomography (CT) was performed before bronchoscopy. Bronchiectasis was assessed by blinded reads, based on the modified Reiff score [4], by two board-certified radiologists from the Japanese Radiological Society with an average experience of 25 years. The Reiff score indicates the degree of bronchial dilatation (tubular, 1; varicose, 2; and cystic, 3) and number of lobes involved. The lingular segment was evaluated as an independent lobe. Scores ranged from 1 to 18, and patients without bronchodilation were assigned a score of 0.

### Identification of the lower respiratory tract-colonizing microbes

To detect the colonizing microorganisms in the LRT, bronchial lavage or intratracheal sputum collected by bronchoscopy was directly cultured using blood with chocolate agar (37 °C, 5% CO_2_), anero Columbia agar (anaerobic chamber), and bromo thymol blue with chromagar (37 °C) for detecting gram-positive, gram-negative, and anaerobic bacteria and fungi.

### Endpoint

The data were censored on March 31, 2019. Pneumonia incidence and pneumonia-free survival, calculated from the date of bronchoscopy to the date of pneumonia diagnosis, were recorded. Patients who did not develop pneumonia or who were lost to follow-up were censored at the date of last contact. Pneumonia was confirmed by the presence of new lung infiltrates and at least one of the following acute respiratory symptoms: cough, sputum production, dyspnea, a body temperature of ≥38.0 °C, abnormal auscultatory findings, and leukocyte counts of >10000 cells/μL or <4000 cells/μL.

### Statistical analysis

The Mann–Whitney U test was used for analyzing continuous variables, and Fisher’s exact test for categorical variables. The cumulative incidence of pneumonia was estimated using the Kaplan–Meier method and log-rank test. Baseline variables with a *P*-value <0.10 (by univariate analysis) and previously reported risk factors for pneumonia in patients with RA were included in the multivariate analysis. The risk of pneumonia was evaluated using Cox proportional-hazards models (CPHM) to determine hazard ratios (HRs) with 95% confidence intervals (95% CIs). A *P*-value <0.05 was considered statistically significant. Analyses were performed using JMP®□ 13 (SAS Institute Inc., Cary, NC, USA).

## Results

### Eligible patients

Among the 228 patients with RA who underwent bronchoscopy during the study period, 107 were excluded from the study, including 71 with active infections (pneumonia, 43; non-tuberculosis mycobacteria, 16; cryptococcosis, 7; aspergillosis, 3; *Pneumocystis*, 1; and other mycosis, 1); 33 with malignancies (lung, 28; renal, esophageal, ovarian, endometrial, and adult T-cell leukemia-lymphoma; 1 each); 1 with a tracheotomy; and 1 with solid organ transplantation. No data were available for 1 patient who was also excluded. Consequently, 121 patients were included in the study (Fig 1).

**Fig 1.**
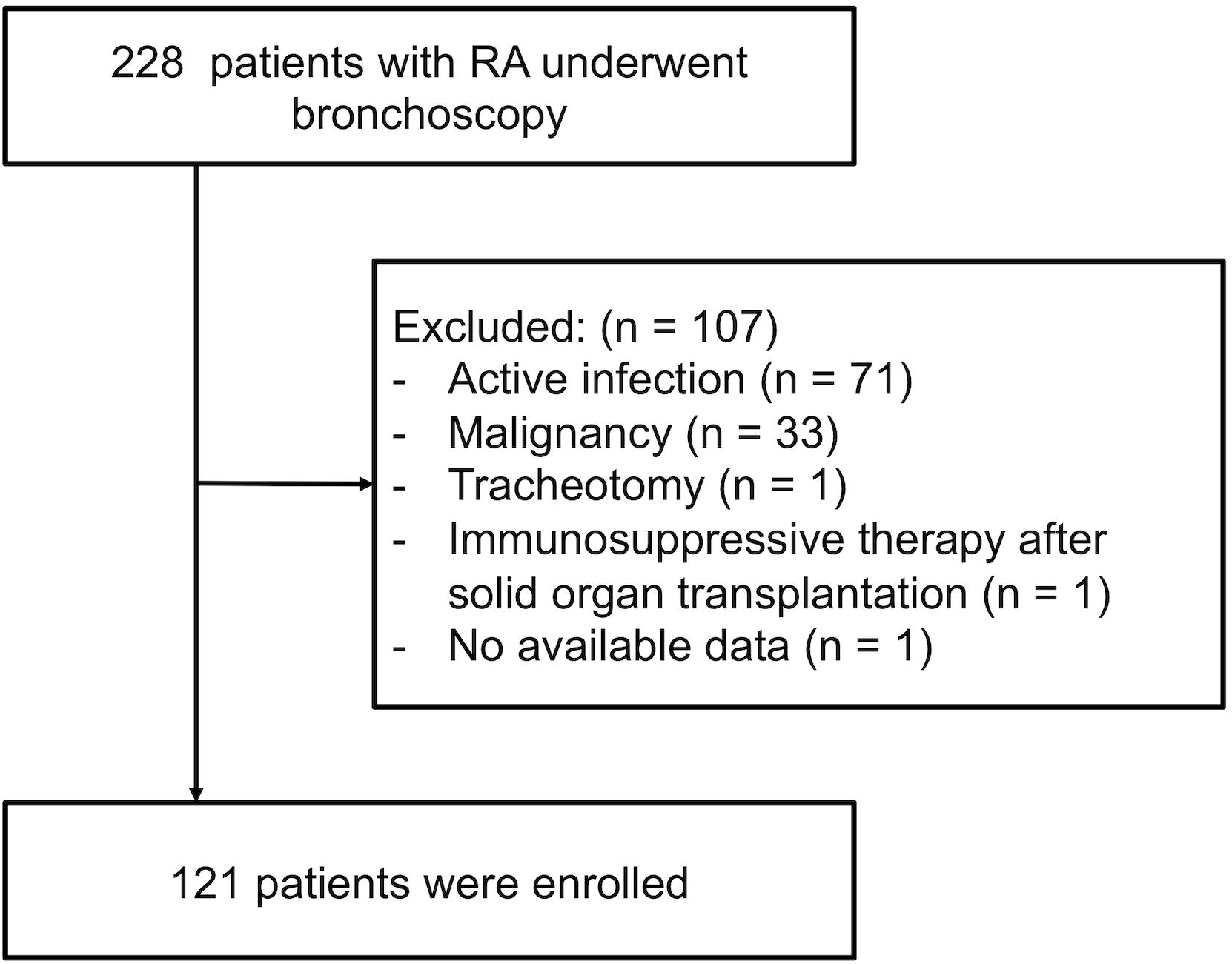
Selection of eligible patients. RA, rheumatoid arthritis

### Lower respiratory tract colonization by microbes

The detected microbes in the LRT samples are listed in Table 1. The LRT samples tested positive in 100 patients (82.6%), including 41 (33.9%) with normal oral flora. Among the frequently isolated pathogens, *P. aeruginosa* (13.2%), *Staphylococcus aureus* (12.4%), and *Haemophilus influenzae* (5.0%) were the most prominent. The other pathogens were *Streptococcus pneumoniae* (1.7%), other streptococci (3.3%), other gram-negative bacilli (5.0%), anaerobic bacteria (1.7%), nontuberculous mycobacteria (4.1%), *Candida* species (3.3%), and other fungi (2.5%).

**Table 1.**
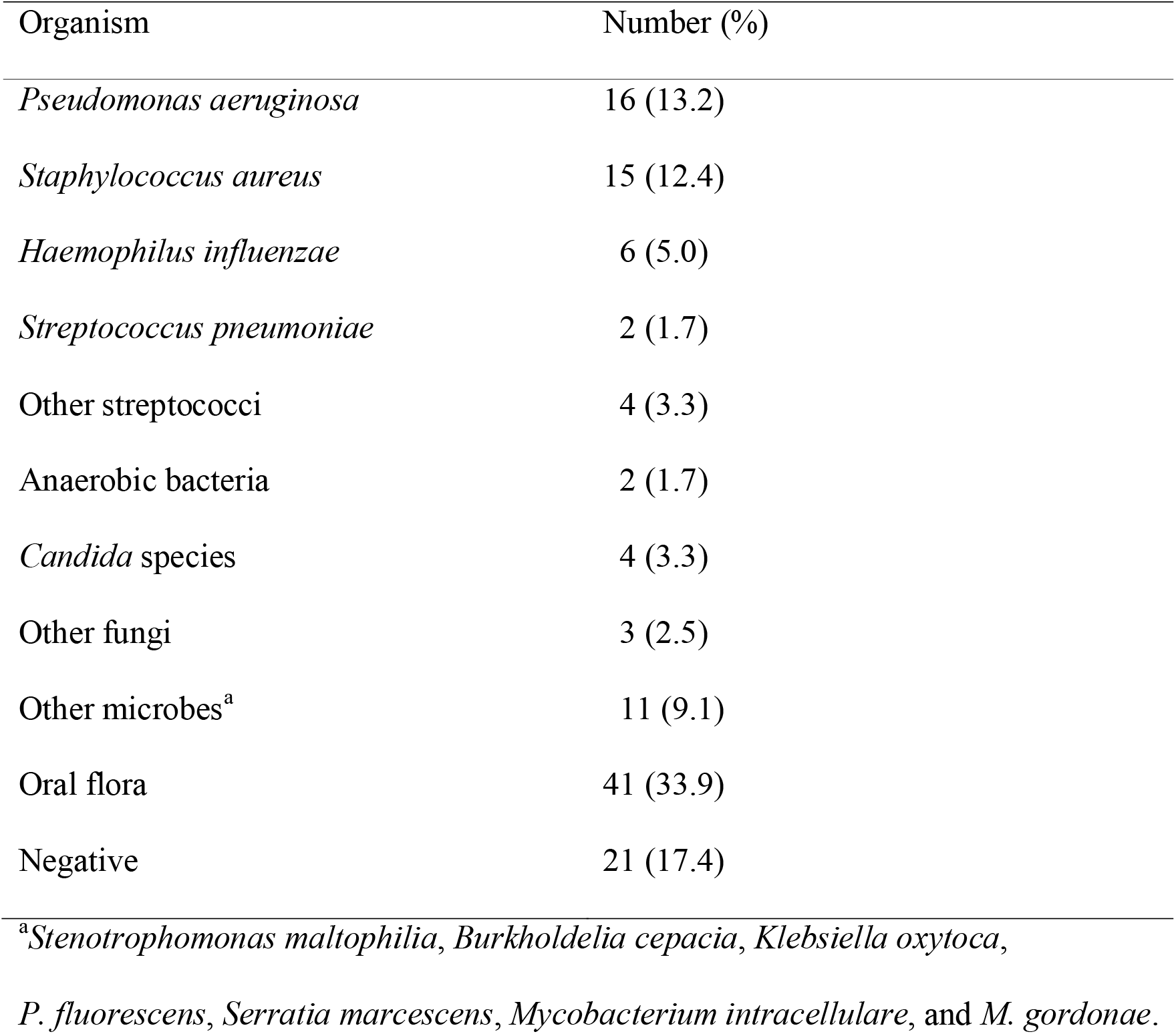
Microorganisms detected by bronchoscopy.

### Patient groups based on the lower respiratory tract-colonizing microbes

Patients with RA were divided into groups depending on the microorganisms isolated from the LRT—*P. aeruginosa* (*Pa* group), *S. aureus* (*Sa* group), and *H. influenzae* (*Hi* group). The others, including those whose samples tested negative (n = 21) or positive for normal oral flora and other pathogens (n = 22), were considered as the control group and were compared with the abovementioned bacteria groups (Table 2).

**Table 2.**
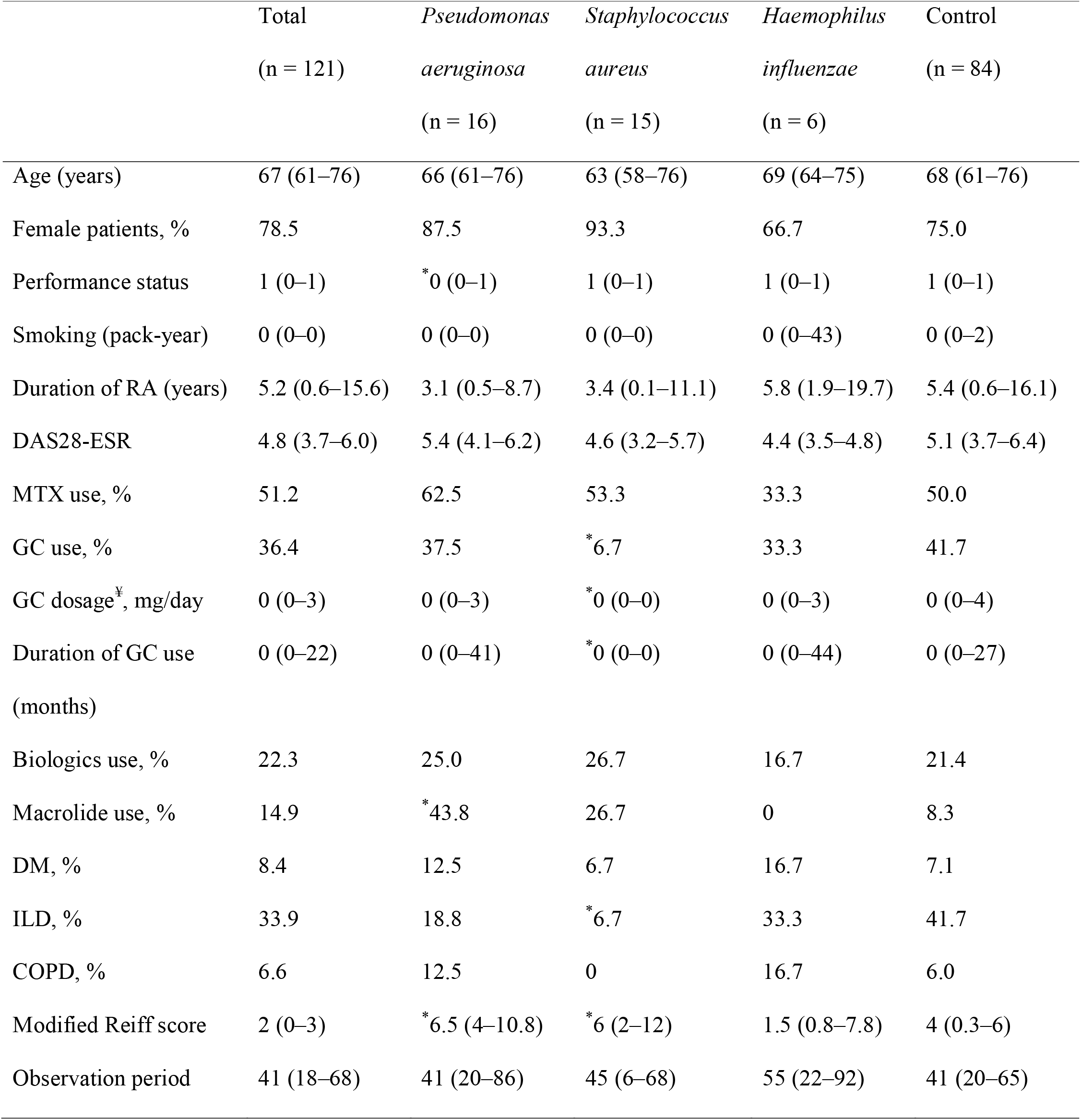
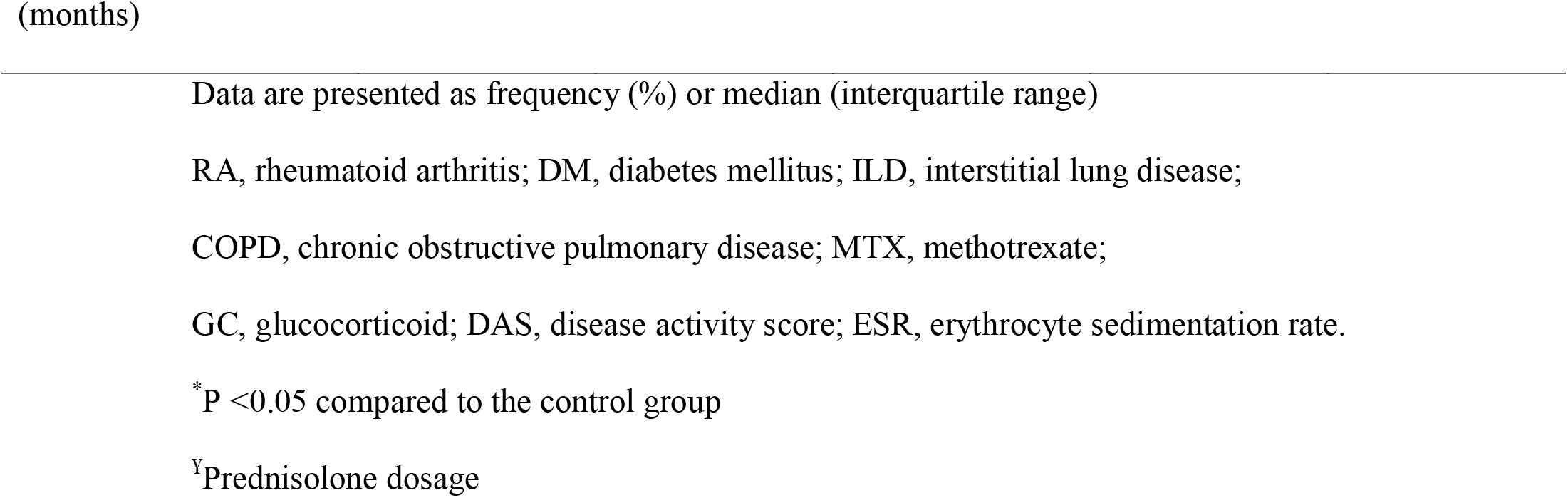
Characteristics of patients with RA, including those whose lower respiratory tract was colonized by each of the three major bacterial species.

|  | Total<br>(n = 121) | *Pseudomonas*<br>*aeruginosa*<br>(n = 16) | *Staphylococcus*<br>*aureus*<br>(n = 15) | *Haemophilus*<br>*influenzae*<br>(n = 6) | Control<br>(n = 84) |
| --- | --- | --- | --- | --- | --- |
| Age (years) | 67 (61–76) | 66 (61–76) | 63 (58–76) | 69 (64–75) | 68 (61–76) |
| Female patients, % | 78.5 | 87.5 | 93.3 | 66.7 | 75.0 |
| Performance status | 1 (0–1) | * 0 (0–1) | 1 (0–1) | 1 (0–1) | 1 (0–1) |
| Smoking (pack-year) | 0 (0–0) | 0 (0–0) | 0 (0–0) | 0 (0–43) | 0 (0–2) |
| Duration of RA (years) | 5.2 (0.6–15.6) | 3.1 (0.5–8.7) | 3.4 (0.1–11.1) | 5.8 (1.9–19.7) | 5.4 (0.6–16.1) |
| DAS28-ESR | 4.8 (3.7–6.0) | 5.4 (4.1–6.2) | 4.6 (3.2–5.7) | 4.4 (3.5–4.8) | 5.1 (3.7–6.4) |
| MTX use, % | 51.2 | 62.5 | 53.3 | 33.3 | 50.0 |
| GC use, % | 36.4 | 37.5 | * 6.7 | 33.3 | 41.7 |
| GC dosage<sup>‡</sup>, mg/day | 0 (0–3) | 0 (0–3) | * 0 (0–0) | 0 (0–3) | 0 (0–4) |
| Duration of GC use<br>(months) | 0 (0–22) | 0 (0–41) | * 0 (0–0) | 0 (0–44) | 0 (0–27) |
| Biologics use, % | 22.3 | 25.0 | 26.7 | 16.7 | 21.4 |
| Macrolide use, % | 14.9 | * 43.8 | 26.7 | 0 | 8.3 |
| DM, % | 8.4 | 12.5 | 6.7 | 16.7 | 7.1 |
| ILD, % | 33.9 | 18.8 | * 6.7 | 33.3 | 41.7 |
| COPD, % | 6.6 | 12.5 | 0 | 16.7 | 6.0 |
| Modified Reiff score | 2 (0–3) | * 6.5 (4–10.8) | * 6 (2–12) | 1.5 (0.8–7.8) | 4 (0.3–6) |
| Observation period | 41 (18–68) | 41 (20–86) | 45 (6–68) | 55 (22–92) | 41 (20–65) |

Data are presented as frequency (%) or median (interquartile range)
RA, rheumatoid arthritis; DM, diabetes mellitus; ILD, interstitial lung disease;
COPD, chronic obstructive pulmonary disease; MTX, methotrexate;
GC, glucocorticoid; DAS, disease activity score; ESR, erythrocyte sedimentation rate.
\* $P < 0.05$ compared to the control group
‡Prednisolone dosage

### Patient backgrounds per group (based on the lower respiratory tract-colonizing microbes)

The age, sex, smoking history, RA duration, and observation period were not different among the groups. The incidence of diabetes mellitus (DM), chronic obstructive pulmonary disease (COPD), and interstitial lung disease (ILD) was relatively low in the *Sa* group. None of the patients in the *Sa* group used glucocorticoids (GCs). Long-term macrolides (≥1 month) were more commonly implemented in the *Pa* group. The *Pa* and *Sa* groups showed a significantly higher degree of bronchiectasis, detected by the modified Reiff score.

### Incidence and hazard ratios of pneumonia

The annual incidence rates of pneumonia per 1000 patients were 100, 62, 132, and 38, in the *Pa, Sa, Hi*, and control groups, respectively. Kaplan–Meier curves are shown in Fig 2. Log-rank tests revealed that the *Pa* group had a significantly higher incidence of pneumonia (*P* = 0.038). The *Hi* group was not analyzed using CPHM since no proportional hazard was established. *S. aureus* colonization was not a statistically significant risk factor for subsequent pneumonia. To compare the *Pa* group with the control group, the HRs were adjusted for age, sex, performance status, DM, ILD, COPD, GCs, and the modified Reiff score using CPHM. *P. aeruginosa* colonization was an independent risk factor for pneumonia in patients with RA (HR, 3.504; 95% CI, 1.153–10.330; Table 3).

**Table 3.** Hazard ratios for pneumonia.

|  | Univariate analysis |  | Multivariate analysis |  |
| --- | --- | --- | --- | --- |
|  | HR (95% CI) | *P*-value | HR (95% CI) | *P*-value |
| Age | 1.039 (0.991–1.094) | 0.119 | 0.999 (0.939–1.067) | 0.971 |
| Female sex | 0.547 (0.203–1.726) | 0.281 | 0.549 (0.139–2.385) | 0.409 |
| Smoking | 1.007 (0.983–1.023) | 0.526 |  |  |
| Performance status | 1.468 (0.780–2.561) | 0.222 | 1.354 (0.612–2.798) | 0.431 |
| DM | 3.860 (0.873–12.228) | 0.071 | 1.714 (0.307–7.951) | 0.515 |
| ILD | 1.794 (0.721–4.527) | 0.201 | 2.554 (0.831–8.169) | 0.102 |
| COPD | 3.547 (0.815–10.929) | 0.084 | 1.624 (0.304–7.647) | 0.546 |
| Duration of RA | 1.022 (0.982–1.057) | 0.264 |  |  |
| MTX | 0.770 (0.308–1.951) | 0.574 |  |  |
| GC use | 1.564 (0.617–3.912) | 0.338 | 1.319 (0.405–4.061) | 0.636 |
| Biologics | 0.824 (0.234–2.284) | 0.728 |  |  |
| Macrolide | 0.799 (0.183–2.457) | 0.719 |  |  |
| DAS28-ESR | 0.946 (0.664–1.356) | 0.760 |  |  |
| Modified Reiff<br>score | 1.106 (0.983–1.247) | 0.093 | 1.135 (0.982–1.311) | 0.084 |
| *Pa* colonization | 2.636 (0.965–6.727) | 0.058 | 3.504 (1.153–10.330) | 0.028 |

HR, hazard ratio; CI, confidence interval; ILD, interstitial lung disease; COPD, chronic obstructive pulmonary disease; RA, rheumatoid arthritis; MTX, methotrexate; GC, glucocorticoid; DAS, disease activity score; ESR, erythrocyte sedimentation rate; *Pa*, *Pseudomonas aeruginosa*

**Fig 2.**
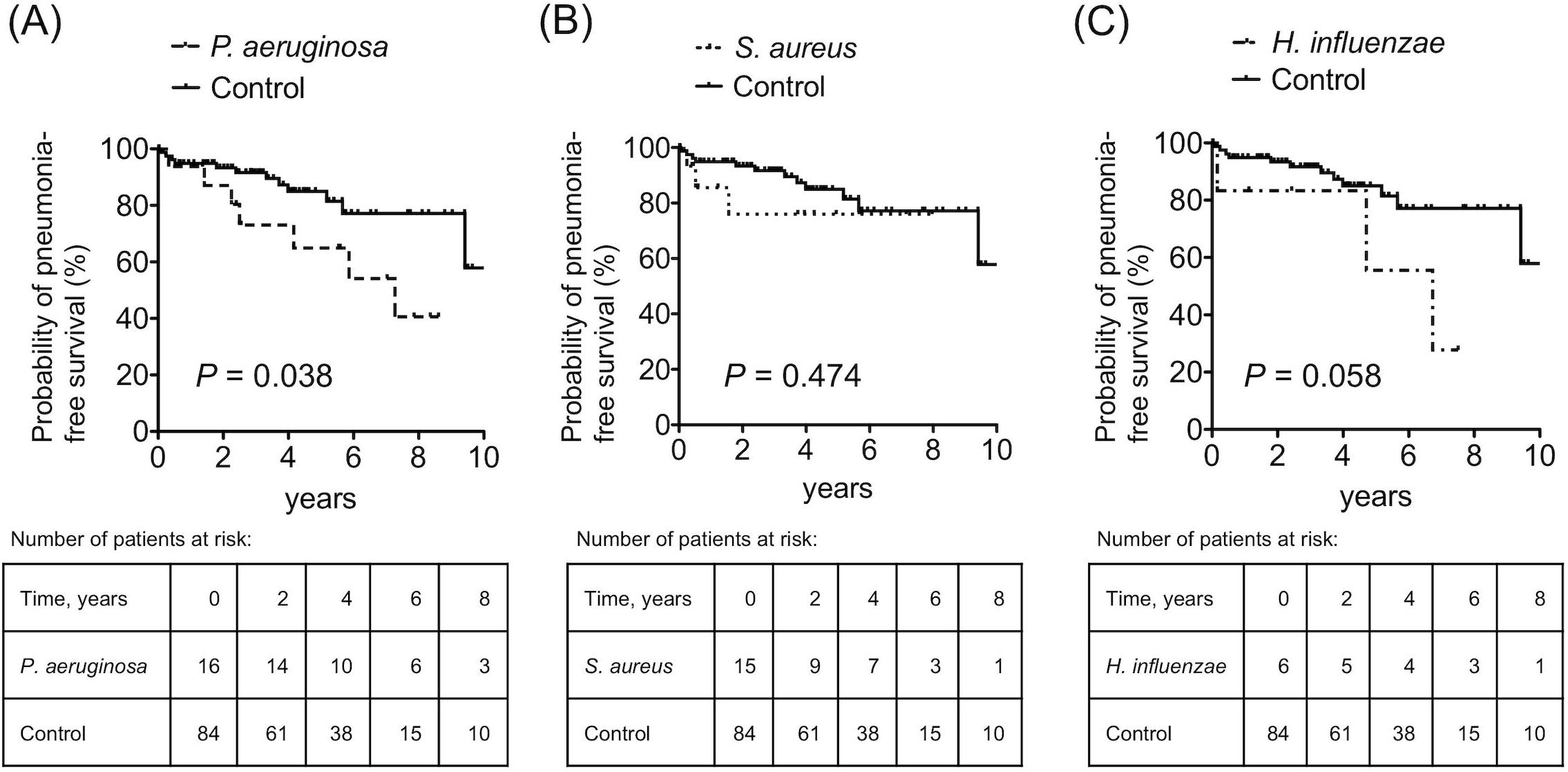
Kaplan–Meier plots and number-at-risk tables for the probability of pneumonia-free survival. Control group: patients with microbes other than *Pseudomonas aeruginosa, Staphylococcus aureus*, and *Haemophilus influenzae*.

## Discussion

We retrospectively analyzed pneumonia in patients with RA based on the microbes that colonized the LRT. *P. aeruginosa, S. aureus*, and *H. influenzae* were the most frequent colonizers, and *P. aeruginosa* colonization was an independent risk factor for subsequent pneumonia in patients with RA. The annual incidence rate of pneumonia per 1000 patients in these three groups was higher than that in the control group, while the frequency of pneumonia in the control group in this study was similar to the frequency in patients with RA in a previous study [7]. To the best of our knowledge, this is the first study that focuses on microbial colonization of the LRT as a risk factor for pneumonia in patients with RA. The strength of this study was the evaluation of microbes by bronchoscopy, minimizing oral bacterial contamination and reflecting LRT-specific flora. Another strength was the novel approach to evaluate whether colonized *P. aeruginosa* was directly correlated with subsequent pneumonia in patients with RA.

*P. aeruginosa* has been reported as one of the major causative microorganisms of pneumonia in patients with RA [1]. The risks of pneumonia in the *Pa* group may have been underestimated in this study due to frequent macrolide use in older patients. Long-term macrolide treatment can reduce pulmonary exacerbation and improve bronchiectasis in non-CF [8, 9]; moreover, macrolides possibly prevent pneumonia in older adults [10]. A relatively higher prevalence of DM and high-degree bronchiectasis in the *Pa* group may also have affected pneumonia incidences.

Conventional culture methods in non-CF patients with bronchiectasis tend to under-recognize *H. influenzae*, compared to *P. aeruginosa* and *S. aureus* [11]. Therefore, the high incidence of pneumonia in the *Hi* group reported herein might be due to the small sample size. Further, most cases of pneumonia in the *Hi* group occurred more than 5 years after colonization, and *P. aeruginosa* was detected in the sputum of two-thirds of the patients who developed pneumonia; thus, microbial substitution by *P. aeruginosa* was possibly related to the incidence of pneumonia in the *Hi* group.

Reportedly, *P. aeruginosa* and *H. influenzae* are the most frequent colonizers of the LRT in patients with non-CF bronchiectasis [12, 13]. They were also the major colonizers and were isolated in cases of bronchiectasis in patients with RA in our study. *S. aureus* colonization with bronchiectasis is associated with CF or allergic bronchopulmonary aspergillosis [13, 14], although neither was seen in our patients.

The present study has several limitations. First, this study was a retrospective study, and the sample size was small. The history of pneumococcal vaccination was unavailable in the medical records, which might have affected the incidence of pneumonia. Second, the detected microorganisms did not reflect the information of the entire LRT. Furthermore, the conventional culture method has a limit when detecting bacteria, and anaerobic bacteria may have been underestimated [15]. Third, only one point was assessed, and it is unclear if microbial substitution occurred after bronchoscopy. To accurately understand the effect of airway microbes on pneumonia in patients with RA, prospective studies with larger samples are required in the future.

## Conclusions

In patients with RA, *P. aeruginosa, S. aureus*, and *H. influenzae* were the three major microbes isolated from the LRT, and the incidence of pneumonia was higher in these patients compared to the control group. *P. aeruginosa* colonization was associated with subsequent pneumonia in patients with RA. Our findings may contribute to the management of patients with RA whose LRT is colonized by *P. aeruginosa* by limiting unnecessary GC use, encouraging pneumococcal vaccination, and selecting antimicrobials that affect *P. aeruginosa* in the empiric treatment of pneumonia.

## Data Availability

None

## Acknowledgments

None.

## References

1. Wakabayashi A, Ishiguro T, Takaku Y, Miyahara Y, Kagiyama N, Takayanagi N. Clinical characteristics and prognostic factors of pneumonia in patients with and without rheumatoid arthritis. PLoS One 2018;13 8: e0201799.

2. Mohd Noor N, Mohd Shahrir MS, Shahid MS, Abdul Manap R, Shahizon Azura AM, Azhar Shah S. Clinical and high resolution computed tomography characteristics of patients with rheumatoid arthritis lung disease. Int J Rheum Dis 2009;12 2: 136–44.

3. Mori S, Koga Y, Sugimoto M. Different risk factors between interstitial lung disease and airway disease in rheumatoid arthritis. Respir Med 2012;106 11: 1591–9.

4. De Soyza A, McDonnell MJ, Goeminne PC, Aliberti S, Lonni S, Davison J et al. Bronchiectasis Rheumatoid Overlap Syndrome Is an Independent Risk Factor for Mortality in Patients With Bronchiectasis. Chest 2017;151 6: 1247–54.

5. Finch S, McDonnell MJ, Abo-Leyah H, Aliberti S, Chalmers JD. A Comprehensive Analysis of the Impact ofPseudomonas aeruginosaColonisation on Prognosis in Adult Bronchiectasis. Annals of the American Thoracic Society 2015;12 11: 1602–11.

6. Aletaha D, Neogi T, Silman AJ, Funovits J, Felson DT, Bingham CO, 3rd et al. 2010 rheumatoid arthritis classification criteria: an American College of Rheumatology/European League Against Rheumatism collaborative initiative. Ann Rheum Dis 2010;69 9: 1580–8.

7. Doran MF, Crowson CS, Pond GR, O’Fallon WM, Gabriel SE. Frequency of infection in patients with rheumatoid arthritis compared with controls: a population-based study. Arthritis Rheum 2002;46 9: 2287–93.

8. Kadota J, Mukae H, Ishii H, Nagata T, Kaida H, Tomono K et al. Long-term efficacy and safety of clarithromycin treatment in patients with diffuse panbronchiolitis. Respiratory Medicine 2003;97 7: 844–50.

9. Rogers GB, Bruce KD, Martin ML, Burr LD, Serisier DJ. The effect of long-term macrolide treatment on respiratory microbiota composition in non-cystic fibrosis bronchiectasis: an analysis from the randomised, double-blind, placebo-controlled BLESS trial. The Lancet Respiratory Medicine 2014;2 12: 988–96.

10. Yoshikawa H, Komiya K, Umeki K, Kadota J-i. Long-Term Macrolide Antibiotic Therapy May Prevent the Development of Pneumonia in the Elderly. Journal of Palliative Medicine 2014;17 7: 749–50.

11. Cox MJ, Turek EM, Hennessy C, Mirza GK, James PL, Coleman M et al. Longitudinal assessment of sputum microbiome by sequencing of the 16S rRNA gene in non-cystic fibrosis bronchiectasis patients. PLoS One 2017;12 2: e0170622.

12. Dimakou K, Triantafillidou C, Toumbis M, Tsikritsaki K, Malagari K, Bakakos P. Non CF-bronchiectasis: Aetiologic approach, clinical, radiological, microbiological and functional profile in 277 patients. Respiratory Medicine 2016;116: 1–7.

13. Pasteur MC, Helliwell SM, Houghton SJ, Webb SC, Foweraker JE, Coulden RA et al. An investigation into causative factors in patients with bronchiectasis. Am J Respir Crit Care Med 2000;162 4 Pt 1: 1277–84.

14. Shah PL, Mawdsley S, Nash K, Cullinan P, Cole PJ, Wilson R. Determinants of chronic infection with Staphylococcus aureus in patients with bronchiectasis. Eur Respir J 1999;14 6: 1340–4.

15. Yamasaki K, Mukae H, Kawanami T, Fukuda K, Noguchi S, Akata K et al. Possible role of anaerobes in the pathogenesis of nontuberculous mycobacterial infection. Respirology 2015;20 5: 758–65.

